# Smoking prevalence and purchasing of menthol cigarettes since the menthol flavour ban in Great Britain: a population-based survey between 2020 and 2023

**DOI:** 10.1101/2023.09.08.23295247

**Authors:** Vera Buss, Harry Tattan-Birch, Sharon Cox, Linda Bauld, Lion Shahab, Jamie Brown

**Affiliations:** Department of Behavioural Science and Health, University College London, UK; SPECTRUM Research Consortium; Usher Institute, College of Medicine and Veterinary Medicine, University of Edinburgh, UK

**Keywords:** Health Policy, Tobacco Smoking, Menthol, Great Britain

## Abstract

**Background:** Menthol cigarettes have been banned in Great Britain (GB) since May 2020. Still, menthol accessories and unlabelled cigarettes perceived as mentholated are available, and people can buy menthol cigarettes overseas or illicitly. This study assessed: trends in smoking menthol cigarettes among all adults and 18-to-24-year-olds in GB between October 2020 and March 2023; trends in and differences between England, Scotland, and Wales during the same period; and purchase sources among people smoking menthol versus non-flavoured cigarettes.

**Methods:** Population-weighted data were from a monthly cross-sectional survey of adults in GB. Among people smoking cigarettes, we calculated the proportion smoking menthol cigarettes across all adults and 18-to-24-year-olds, and prevalence ratios (PR) between first and last quarter. We also calculated proportions of people smoking menthol/non-flavoured cigarettes by purchase source (including illicit sources).

**Results:** In the first quarter, 16.2% of adults smoking cigarettes reported menthol cigarette smoking with little to no decline throughout the study (PR=0.85, 0.71-1.01), while it declined slightly among 18-to-24-year-olds (PR=0.75, 0.63-0.89). The prevalence of menthol cigarette smoking fell by two-thirds in Wales (PR=0.36, 0.19-0.62) but remained relatively stable in England (PR=0.88, 0.72-1.06) and Scotland (PR=0.94, 0.59-1.53). The main purchasing sources were licit (93.9%), 14.8% reported illicit sources and 11.5% cross-border purchases, without notable differences from people smoking non-flavoured cigarettes.

**Conclusions:** Roughly one million adults in GB still smoke menthol cigarettes and, with the exception of Wales, there were no noteworthy changes in the post-ban period. There was no indication that this was driven by illicit purchases.

**What is already known on this topic:** Tobacco companies have used various loopholes in the legislation to circumvent the menthol cigarette ban in Great Britain and, in general, some people tend to migrate towards illicit purchases when their product is banned.

**What this study adds:** Despite the ban, menthol cigarettes have remained popular among adults who smoke in Great Britain, with roughly one in seven reporting smoking menthol cigarettes. Between October 2020 and March 2023, there was no noteworthy change in menthol cigarette smoking prevalence in the overall British adult population, but there was a sharp decline among the Welsh population.

**How this study might affect research, practice or policy:** Since the majority of people who reported menthol cigarette smoking purchased cigarettes through licit sources, it might indicate that most of them either use accessories to add menthol flavour to their cigarettes or they purchase cigarette brands that are perceived to contain menthol flavouring without being labelled as such. If the aim is to reduce menthol cigarette smoking prevalence to nearly zero, policymakers in Great Britain should consider closing loopholes in the current legislation, such as prohibiting all menthol and its analogues and derivatives in all tobacco-related products, including accessories.

## INTRODUCTION

Factory-made and roll-your-own tobacco with characterising flavours (that alter the smell and taste of the product) have been banned in the United Kingdom (UK) and the European Union since May 2020 [1, 2]. Menthol is the most common cigarette flavour and menthol cigarettes are particularly popular among youth (aged 12-17) and young adults (aged 18-24) because menthol reduces negative sensory characteristics associated with smoking, and menthol cigarettes are misperceived as less harmful [3–5]. Previous research showed that prevalence of menthol cigarette smoking has remained high in the year after the ban [6, 7]. It is important for policymakers to know whether the relatively high prevalence has persisted, and if so, what the main drivers are.

As one of the main intentions of this legislation was to reduce smoking uptake among youth, other tobacco products containing menthol such as cigars were exempted from the ban due to low sales volumes or consumption among young people [2]. It is assumed that a menthol cigarette ban would lead to a decrease in smoking prevalence as fewer young people would start smoking [5, 8]. Another rationale for the ban is that people who smoke menthol cigarettes will be more likely to quit when they can no longer purchase menthol cigarettes than to switch to non-flavoured cigarettes [5, 8–10].

There are several reasons why people in the UK may continue to smoke menthol cigarettes despite the ban. First, it is possible to buy factory-made cigarettes or roll-your-own tobacco with menthol flavour in countries without a ban and bring them back to the UK either within the legal limits for personal use or through illicit means. Second, people can purchase menthol accessories, such as filters or capsules inserted in a hole in filters of factory-made cigarettes, infusion cards for cigarette packs to spread menthol aroma and flavour, or menthol-flavoured filters for use with roll-your-own tobacco [11]. These accessories are not covered by the ban and some of them seem to have been placed on the UK market in direct response to the ban [11, 12]. Another tactic that the tobacco industry used to circumvent the ban is to produce cigarettes that may be perceived as mentholated, while the manufacturers claim that the flavours are not characterising and are therefore allowed [13]. For example, some menthol cigarette products by Japan Tobacco International were rebranded under a “dual” range, such as Benson & Hedges Dual, and have been accused of still containing menthol [13, 14].

Early results from England after the ban showed a decline in menthol cigarette smoking among young people aged 16 to 19 years, from 12.1% of young people smoking in the past 30 days in February 2020 before the ban was implemented to 3.0% in August 2020 after the implementation [6]. Figures referring to only those who smoked on at least 20 out of the last 30 days showed a decline from 11.1% to 2.0% in the same period [6]. Another study from England found that among current smokers, 15.7% reported menthol cigarette smoking between July 2020 and June 2021 with a decline from April 2021 onwards [7]. Among 16-to-24-year-olds currently smoking, the prevalence was 25.2%. The study also found that among those reporting menthol cigarette smoking, the percentage of young people, women, and people with professional or managerial occupations was higher than among those reporting smoking other cigarettes [7].

The present study aimed to provide an update on menthol cigarette smoking prevalence to assess whether it has continued to decline since June 2021. Further, it included data from all three nations in Great Britain to identify potential differences between England, Scotland, and Wales. While the menthol ban applies to all three nations, Scotland differs from the other two nations in that the government prohibits the display of any tobacco and smoking related products in shops [15, 16], which could mean people living in Scotland are less aware of the availability of menthol-flavoured tobacco accessories. In England and Wales, tobacco accessories can be displayed at the point of sale [15]. The study also evaluated where people who stated that they smoked menthol cigarettes purchased them, to understand purchase patterns (i.e., licit, illicit, or cross-border).

The research questions were: (i) Has the prevalence of smoking menthol cigarettes among all adults who smoke cigarettes and specifically among young adults (18-24 years) smoking cigarettes in Great Britain changed between October 2020 and March 2023? (ii) Were there differences in the change in prevalence of smoking menthol cigarettes as a proportion of all adults who smoke cigarettes between England, Scotland, and Wales? (iii) Where did people who smoke menthol cigarettes purchase tobacco products in Great Britain between October 2020 and March 2023, were these illicit or cross-border purchases, and did the sources of purchase differ from those who smoked non-flavoured cigarettes?

## METHODS

### Study design

Data for this study were drawn from the Smoking Toolkit Study, an ongoing monthly population-based survey including demographic and smoking-related questions [17]. This study includes data collected between October 2020 and March 2023 from England, Scotland, and Wales. Prior to the analysis, the study protocol was published on the Open Science Framework (https://osf.io/s8mjr/). The manuscript followed the Strengthening the reporting of observational studies in epidemiology (STROBE) statement [18]. Data collection was conducted by a market research company using a combination of random location and quota sampling. Anonymised data were provided to the research team. The data collection method changed in March 2020 from face-to-face to telephone surveys. Studies showed similar results when comparing the two data collection methods [19–21].

### Outcome variables and covariates

The primary outcome measures were the prevalence of menthol cigarette smoking as a proportion of all adults who smoke and specifically young adults smoking (18-24 years) and, for each purchasing source, the proportion of individuals smoking menthol cigarettes who states that they purchased cigarettes through this source. First, participants were grouped according to whether they currently smoked cigarettes. Then, they were further classified based on whether they smoked menthol cigarettes (or for sensitivity analysis, any flavoured cigarettes). All variables are listed in Table 1 and are based on self-report.

**Table 1:**
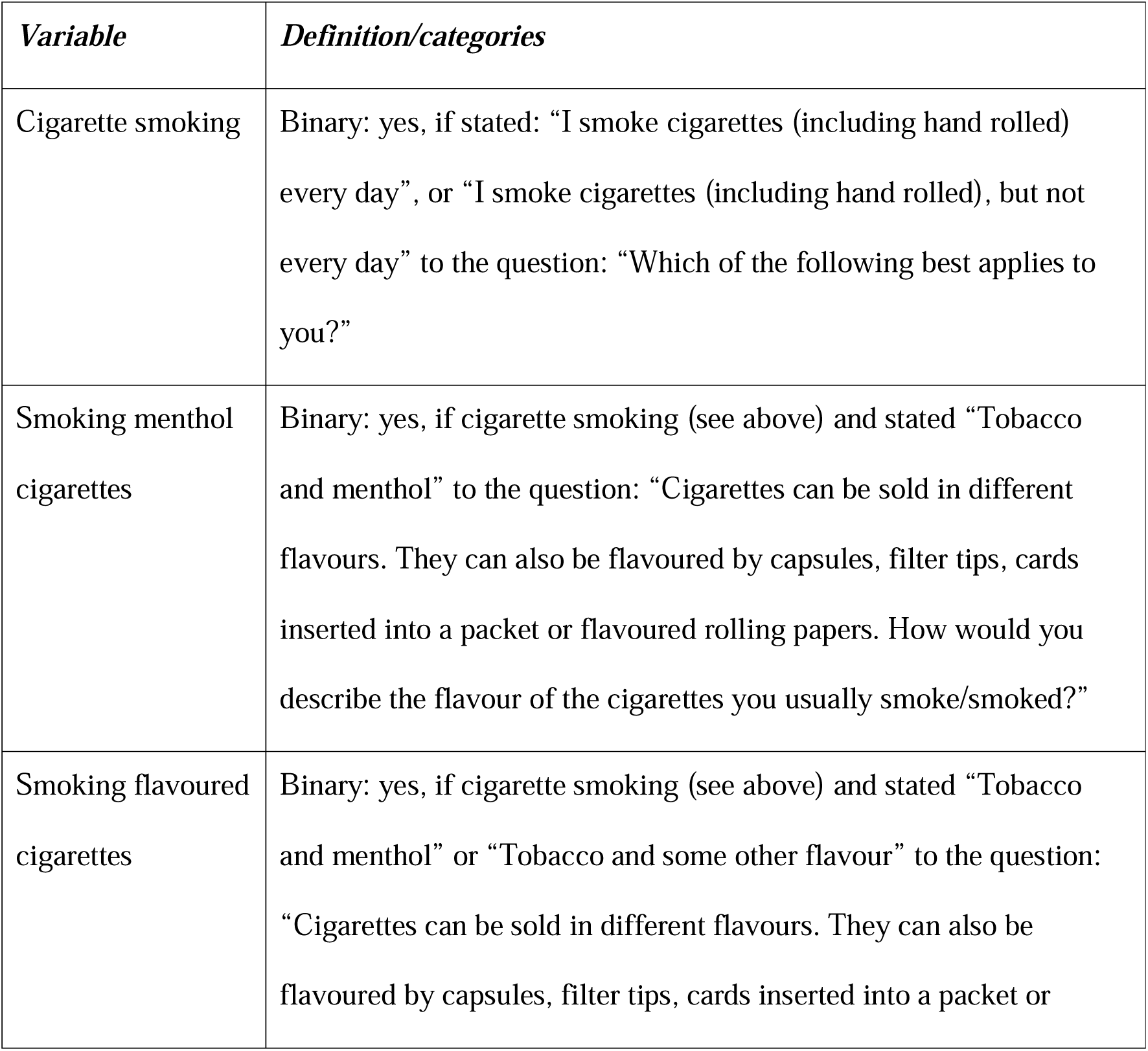
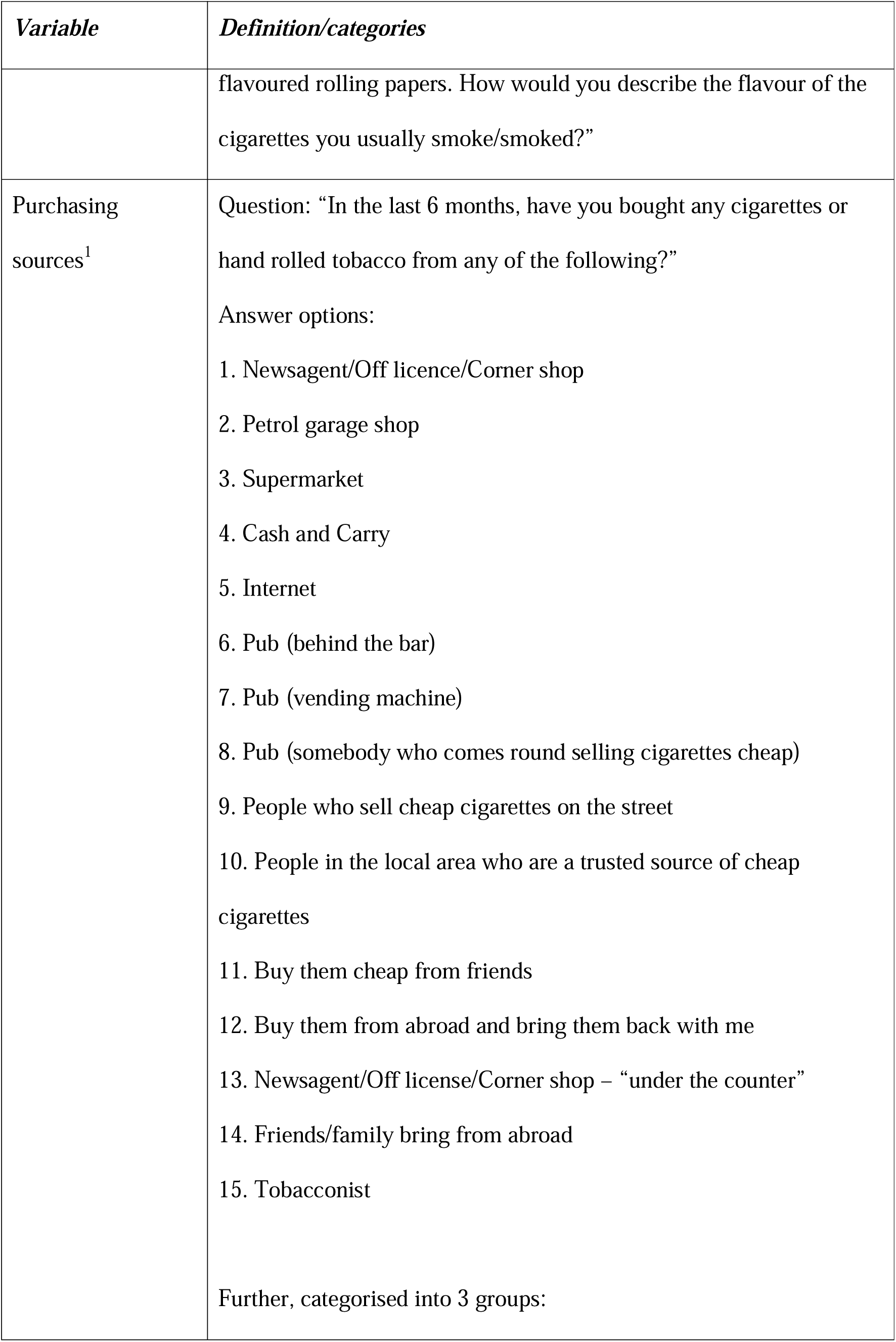
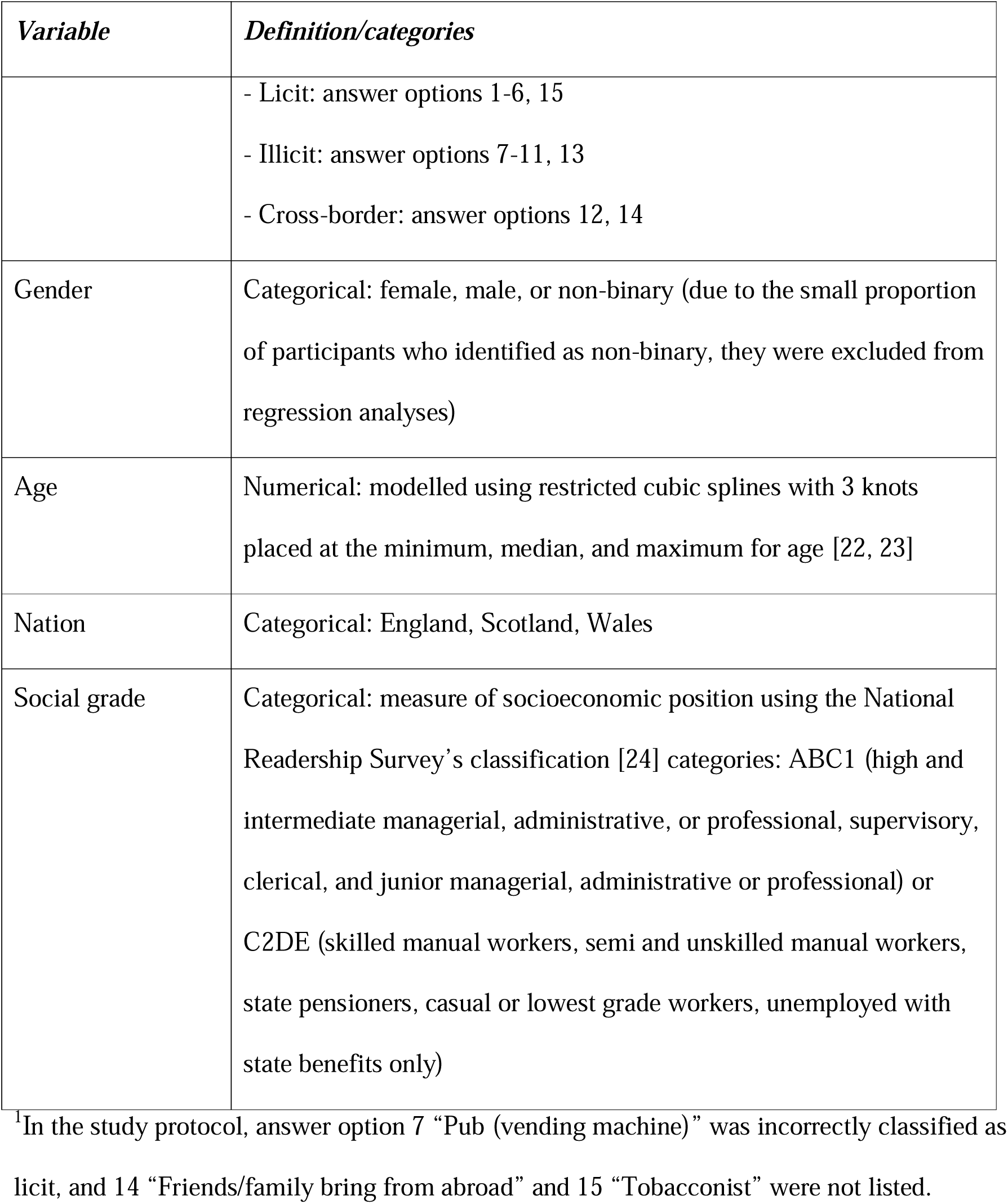
Variables used in analyses, their definitions, and categories if appliable.

### Analysis

The analysis was conducted in RStudio (version 2022.07.2, R version 4.2.1). For each variable included in the analysis, the number and percent of missing values are reported in the supplement (Table S1). All values that the interviewer noted as “Don’t know” or “Refused” were assumed to be missing. The study is based on a complete case analysis. For the first research question, the weighted prevalence of smoking menthol cigarettes as a proportion of all adults in Great Britain who smoke cigarettes was assessed for each quarter between October 2020 and March 2023. Prevalence was reported for quarters rather than months due to small samples in 18-to-24-year-olds, in Scotland and in Wales. Data were weighted using raking to match the population of Great Britain [25]. The weighted prevalence was plotted over time. Further, a logistic regression model with a restricted cubic spline function (3 knots at beginning, middle, and end of time series) was fitted to assess weighted prevalence as a proportion of people smoking menthol cigarettes over time. Prevalence ratios (PR) and corresponding 95% compatibility intervals (CI, using bootstrapping with n=2000 replicates (Cole et al., 2020; Efron & Tibshirani, 1994; Hawkins & Samuels, 2021; Rafi & Greenland, 2020)) were calculated for the change in prevalence comparing the first quarter (Q4 2020) to the last quarter (Q1 2023). Additionally, the weighted prevalence was assessed for young adults aged 18 to 24 years in Great Britain. For the second research question, the weighted prevalence of smoking menthol cigarettes as a proportion of those who smoke cigarettes was assessed for each nation (England, Scotland, and Wales) over time (in quarters). The weighted prevalence of smoking menthol cigarettes was plotted over time for each nation and PRs and corresponding 95% CIs (using bootstrapping with n=2000 replicates) calculated for the change in prevalence in each nation across the study period. Additionally, logistic regression including an interaction term between time and nation was used to calculate the PR ratio (unadjusted and adjusted for age, gender, social grade) of smoking menthol cigarettes among those who smoke cigarettes in Scotland and Wales compared to England (i.e., dividing the PR of Scotland or Wales, respectively, by the PR of England). Corresponding 95% CIs were computed using bootstrapping (n=2000 replicates).

In England, the question about purchasing sources of cigarettes was only asked once per quarter since April 2022 (i.e., in April 2022, July 2022, October 2022, and January 2023). Therefore, months in which data were only collected in Scotland and Wales were excluded from the analysis for the third research question. The weighted proportion of people who smoked menthol cigarettes stating that they purchased cigarettes by the various sources (not mutually exclusive) was computed for the entire study period (excluding the above-mentioned months). Further, weighted proportions of licit, illicit, and cross-border purchases were assessed. The weighted proportion for each purchase source was compared between those who smoked menthol cigarettes and those who did not smoke flavoured cigarettes using Chi-square statistics and Cramer’s V as a measure of the effect size (following the interpretation by Cohen ^26^, categorising Cramer’s V into small, medium, and large effect sizes). Further, purchase sources were compared between nations. In sensitivity analyses, the research questions were assessed including all people who stated that they smoked flavoured cigarettes (menthol or some other flavour) instead of just those who stated that they smoked menthol cigarettes. Further, the prevalence of smoking menthol cigarettes among all adults and specifically young adults in all of Great Britain and the prevalence of smoking menthol cigarettes among all adults separately in the three nations were assessed.

## RESULTS

For 66,868 (98.7%) out of a total of 67,746 participants, complete data were available on all relevant variables, excluding purchasing sources (data on purchasing sources only available for 6,757 out of 9,195 participants who smoked cigarettes due to the months without data collection for this variable; 191 out of 6,757 (2.9%) with missing values). Among these participants, 9,773 (14.6%) smoked cigarettes (see Table 2, unweighted data in supplementary Table S2). The median age was 49 years (interquartile range: 33-63). There were 7,660 participants aged between 18 and 24 years, of which 1,536 (20.1%) smoked cigarettes.

**Table 2:** Characteristics of survey respondents (N = 66,868; data weighted).

| *Characteristic* | *All* | *England*<br>(*n* = 57,469) | *Scotland*<br>(*n* = 5,832) | *Wales*<br>(*n* = 3,264) |
| --- | --- | --- | --- | --- |
| Age, median (IQR) | 49 (33-63) | 48 (32-63) | 52 (36-65) | 55 (40-69) |
| Gender, n (%) |  |  |  |  |
| Female | 32427 (48.6) | 28054 (48.7) | 2801 (47.9) | 1572 (48) |
| Male | 33918 (50.8) | 29258 (50.8) | 2996 (51.3) | 1664 (50.8) |
| Non-binary | 400 (0.6) | 316 (0.5) | 45 (0.8) | 39 (1.2) |
| Social grade, n (%) |  |  |  |  |
| ABC | 37341 (55.9) | 32416 (56.3) | 3188 (54.6) | 1738 (53.1) |
| C2DE | 29404 (44.1) | 25212 (43.7) | 2654 (45.4) | 1537 (46.9) |
| Cigarette smoking, n<br>(%) | 9773 (14.6) | 8480 (14.7) | 817 (14) | 477 (14.6) |

The prevalence of menthol cigarette smoking in all adults and those aged 18-to-24-years who smoked cigarettes in Great Britain, and adults in England, Scotland, and Wales are listed by quarter in the supplement (weighted data in Table S3, unweighted data in Table S4). Figure 1 shows that the prevalence of menthol cigarette smoking was relatively stable over time among all adults, at 16.2% in the first quarter (Q4 2020) and 13.7% in the final quarter (Q1 2023; PR = 0.85, 95% CI: 0.71-1.01). Assuming an adult population of 52 million adults in GB [27], this results in roughly one million adults smoking menthol cigarettes in the first quarter of 2023. Among 18-to-24-year-olds, the prevalence declined by a quarter from 25.7% to 19.4% (PR=0.75, 95% CI: 0.63-0.89).

**Figure 1:**
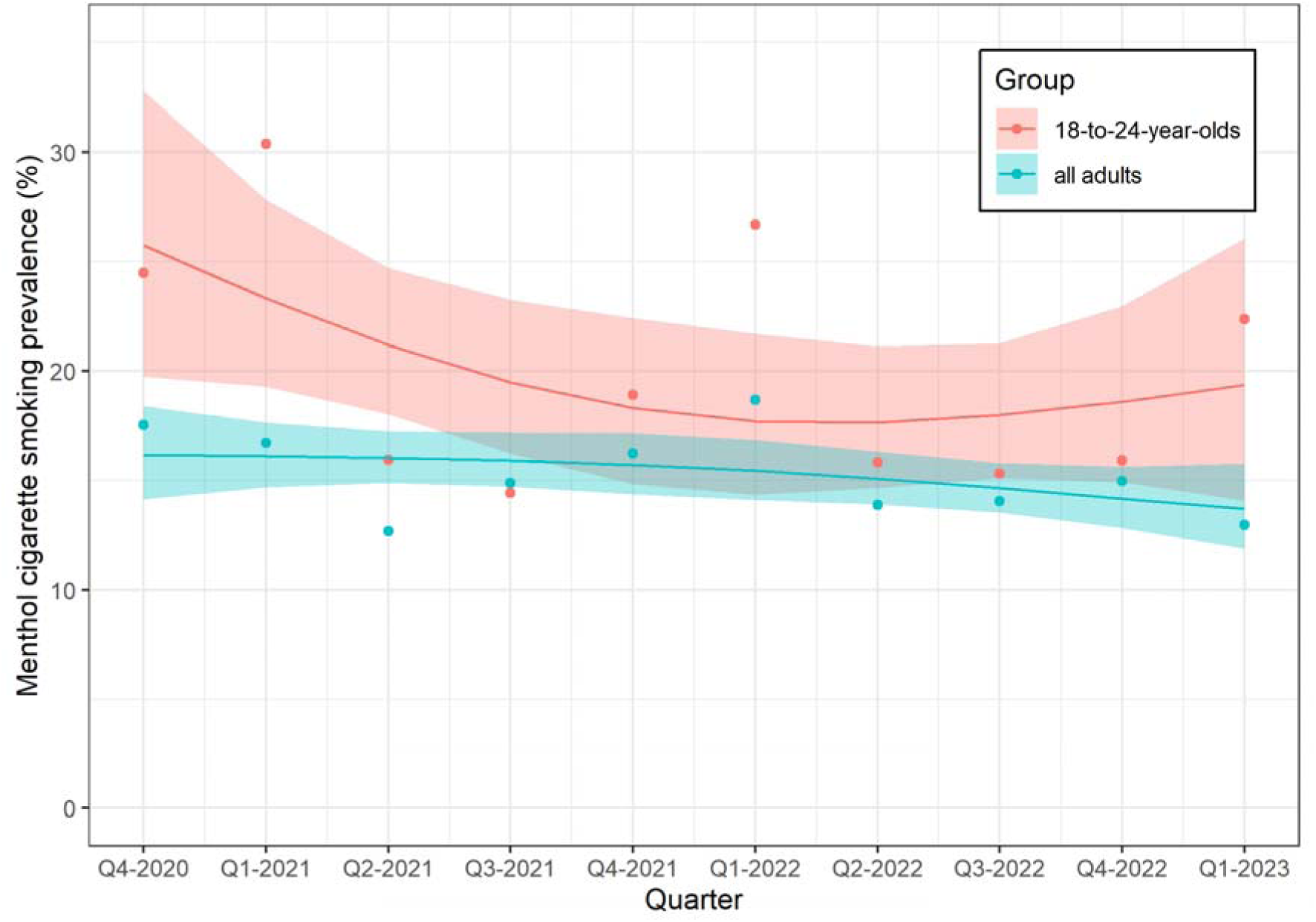
Weighted prevalence of smoking menthol cigarettes among all adults who smoke cigarettes and those aged 18-24 years in Great Britain over time. Lines and shaded bands represent point estimates and 95% compatibility intervals, respectively, from logistic regression with time modelled with restricted cubic splines (3 knots). The points represent unmodelled data.

Figure 2 shows that the prevalence of menthol cigarette smoking was relatively stable over time in England, at 16.2% in the first quarter (Q4 2020) and 14.2% in the final quarter (Q1 2023; PR = 0.88, 95% CI: 0.72-1.06), and Scotland, at 12.0% in the first quarter and 11.3% in the final quarter (PR=0.94, 95% CI: 0.59-1.53). In Wales, the prevalence decreased by almost two-thirds over time, from 22.5% in the first quarter to 8.1% in the last quarter (PR=0.36, 95% CI: 0.19-0.62). There was initially a higher prevalence in Wales than England (modelled estimates for Q4 2020: 22.5% vs. 16.2%) but a lower prevalence by 2023 (modelled estimates for Q1 2023: 8.1% vs. 14.2%; unadjusted PR ratio=0.41, 95% CI: 0.21-0.75; adjusted PR ratio=0.34, 95% CI: 0.21-0.83). Conversely, Scotland had a lower prevalence than in England throughout the whole period (modelled estimates for Q4 2020: 12.0% vs. 16.2, and for Q1 2023: 11.3% vs. 14.2%; unadjusted PR ratio=1.07, 95% CI: 0.64-1.80; adjusted PR ratio=1.21, 95% CI: 0.69-2.07, Figure 3).

**Figure 2:**
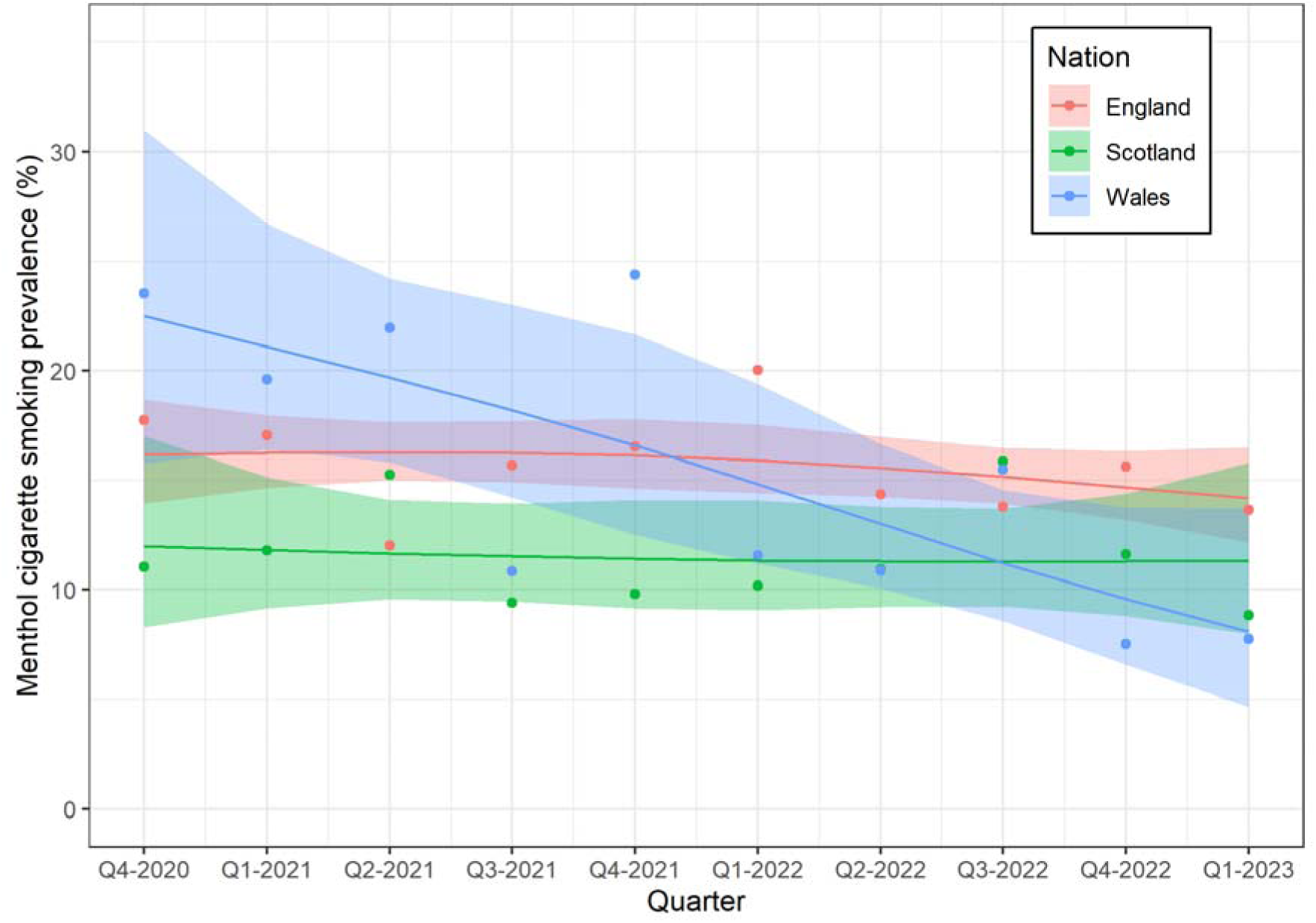
Weighted prevalence of smoking menthol cigarettes among adults who smoke in England, Scotland, and Wales over time. Lines and shaded bands represent point estimates and 95% compatibility intervals, respectively, from logistic regression with time modelled with restricted cubic splines (3 knots). The points represent unmodelled data.

**Figure 3:**
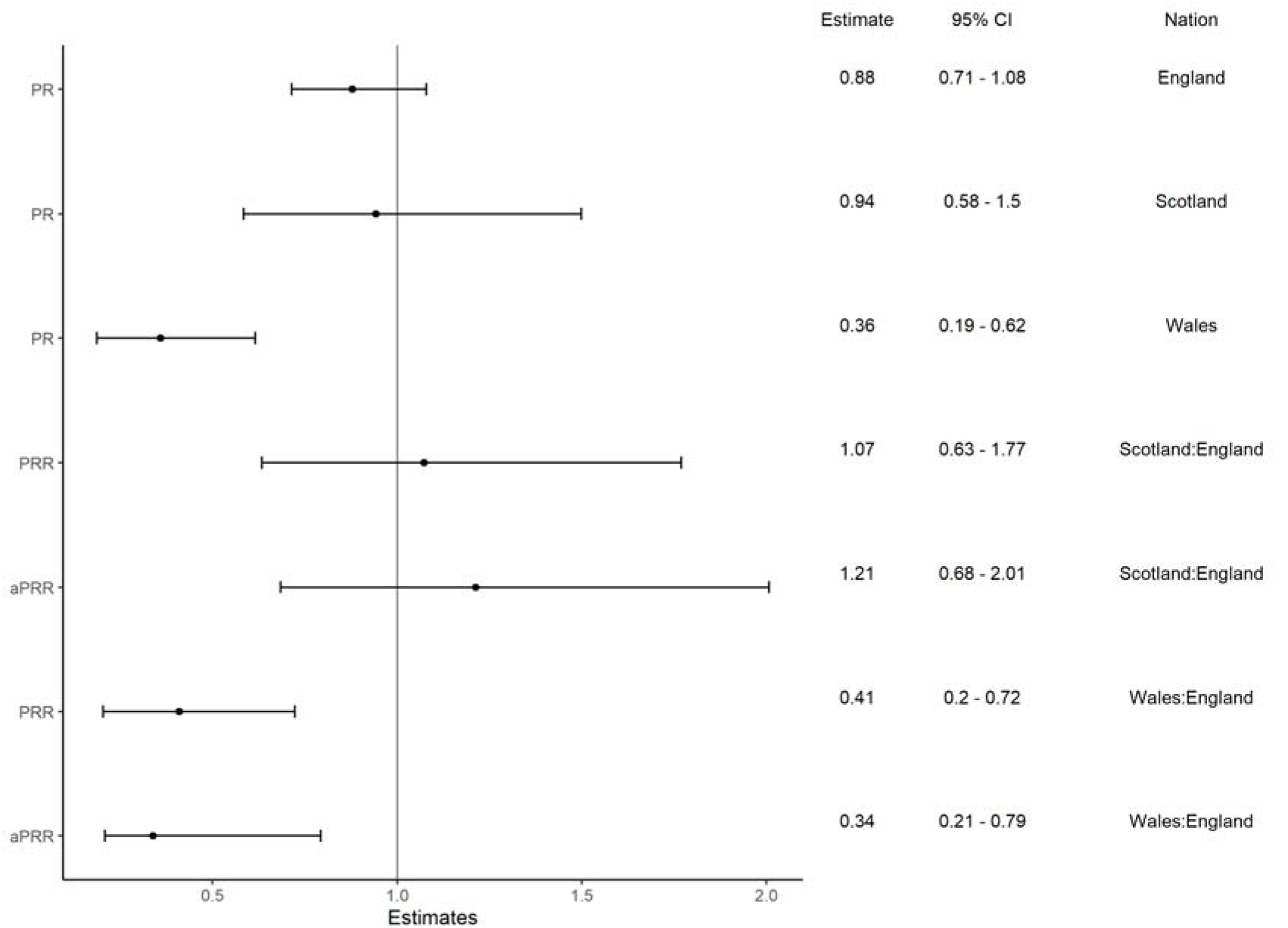
Prevalence ratios comparing menthol smoking among people who smoked in Q4 2020 (reference) to Q1 2023 by nation, and prevalence ratio ratios comparing prevalence ratios between nations. Abbreviations: aPRR, adjusted prevalence ratio ratio (adjusted for age, gender, and social grade using the median (49 years) and the most common category (men, ABC1) as reference); CI, compatibility interval; PR, prevalence ratio; PRR, prevalence ratio ratio.

### Sources of purchasing

Table 4 shows the sources of purchase among those who smoked menthol cigarettes compared to those only smoking non-flavoured cigarettes (unweighted data in supplement, Table S5). The main sources were newsagents/off license/corner shops and supermarkets. There were no noteworthy differences between the two groups. Most participants reported purchasing through licit sources (for menthol cigarette smoking: 93.9%, 95% CI: 92.2-95.5; for non-flavoured cigarette smoking: 93.5%, 95% CI: 92.7-94.2). Illicit sources of purchase were reported by 14.8% (95% CI: 12.2-17.3) of those smoking menthol cigarettes and 12.5% (95% CI: 11.5-13.5) of those smoking only non-flavoured cigarettes. Cross-border purchases were reported by 11.5% (95% CI: 9.2-13.8%) of participants smoking menthol cigarettes and 9.9% (95% CI: 9.0-10.8) of participants smoking only non-flavoured cigarettes.

**Table 3:** Sources of cigarette purchases (not mutually exclusive) among those who smoke menthol cigarettes or non-flavoured cigarettes (n=6621, data weighted).

| *Source of purchase* | *Among people smoking menthol cigarettes, % (95% CI)* | *Among people smoking non-flavoured cigarettes, % (95% CI)* | *P-value<sup>1</sup>* |
| --- | --- | --- | --- |
| Newsagent\Off licence\Corner shop | 72.8 (69.7-75.8) | 69.7 (68.3-71.1) | 0.047 |
| Petrol garage shop | 43.0 (39.6-46.5) | 40.9 (39.4-42.4) | 0.193 |
| Supermarket | 72.4 (69.3-75.5) | 72.2 (70.8-73.5) | 0.885 |
| Cash and Carry | 6.2 (4.4-8.0) | 5.3 (4.6-6.0) | 0.237 |
| Internet | 2.3 (1.3-3.4) | 2.7 (2.2-3.2) | 0.481 |
| Bar in pub | 2.1 (1.0-3.2) | 1.3 (0.9-1.6) | 0.036 |
| Other sources | 0.5 (0.0-1.1) | 1.0 (0.7-1.3) | 0.186 |
| **Illicit** |  |  |  |
| Newsagent\Off license\Corner shop – “under the counter” | 8.2 (6.2-10.2) | 6.0 (5.3-6.8) | 0.009 |
| Friends | 5.9 (4.2-7.6) | 5.6 (4.8-6.3) | 0.683 |
| Trusted local | 3.8 (2.4-5.2) | 3.5 (2.9-4.1) | 0.638 |
| Person in pub | 2.9 (1.6-4.1) | 1.8 (1.4-2.3) | 0.027 |
| Person on the street | 2.3 (1.2-3.3) | 2.3 (1.8-2.8) | 0.952 |
| Vending machine pub | 1.2 (0.4-2.1) | 0.9 (0.6-1.2) | 0.292 |
| **Cross-border** |  |  |  |
| Buy them from abroad and bring them back | 11.3 (9.0-13.5) | 9.7 (8.9-10.6) | 0.136 |
| Friends/family bring from abroad | 0.3 (0.1-0.7) | 0.1 (0.0-0.2) | 0.244 |

<sup>1</sup>All values for Cramer's V were $\leq 0.10$ .

**Table 4:** Sources of cigarette purchases (not mutually exclusive) among those who smoke menthol cigarettes by nation (n=6621, data weighted).

| *Source of purchase* | *Among people smoking menthol cigarettes in England, % (95% CI)* | *Among people smoking menthol cigarettes in Scotland, % (95% CI)* | *Among people smoking menthol cigarettes in Wales, % (95% CI)* | *P-value<sup>1</sup>* |
| --- | --- | --- | --- | --- |
| Newsagent/Off licence/Corner shop | 73.1 (69.7-76.5) | 69.6 (61.5-77.7) | 70.9 (61.1-80.7) | 0.802 |

| *Source of purchase* | *Among people*<br>*smoking menthol*<br>*cigarettes in*<br>*England,*<br>*% (95% CI)* | *Among people*<br>*smoking menthol*<br>*cigarettes in*<br>*Scotland,*<br>*% (95% CI)* | *Among people*<br>*smoking menthol*<br>*cigarettes in*<br>*Wales,*<br>*% (95% CI)* | *P-*<br>*value<sup>1</sup>* |
| --- | --- | --- | --- | --- |
| Petrol garage shop | 42.8 (39.0-46.6) | 40.2 (31.5-48.8) | 50.1 (39.4-60.9) | 0.521 |
| Supermarket | 72.3 (68.9-75.8) | 70.0 (61.7-78.3) | 75.8 (66.6-85.0) | 0.783 |
| Cash and Carry | 6.5 (4.5-8.5) | 5.4 (1.8-9.0) | 2.9 (0.0-5.7) | 0.554 |
| Internet | 2.3 (1.1-3.5) | 3.5 (0.7-6.4) | 1.6 (0.0-3.8) | 0.776 |
| Bar in pub | 2.3 (1.1-3.5) | 1.5 (0.0-4.0) | 0.7 (0.0-2.1) | 0.699 |
| Other sources | 0.5 (0.0-1.1) | 0.4 (0.0-1.3) | 0.0 (0.0-0.0) | 0.846 |
| **Illicit** |  |  |  |  |
| Newsagent/Off<br>license/Corner shop –<br>“under the counter” | 8.4 (6.2-10.6) | 3.0 (0.4-5.6) | 10.9 (3.9-17.8) | 0.256 |
| Friends | 6.1 (4.2-8.0) | 3.4 (0.0-7.3) | 5.3 (1.2-9.4) | 0.687 |
| Trusted local | 4.0 (2.5-5.5) | 0.0 (0.0-0.0) | 5.0 (0.2-9.9) | 0.260 |
| Person in pub | 2.9 (1.5-4.3) | 2.9 (0.0-6.4) | 1.9 (0.0-5.6) | 0.915 |
| Person on the street | 2.2 (1.1-3.3) | 3.6 (0.0-7.7) | 2.5 (0.0-6.4) | 0.772 |
| Vending machine pub | 1.3 (0.4-2.2) | 1.3 (0.0-3.8) | 0.0 (0.0-0.0) | 0.708 |
| **Cross-border** |  |  |  |  |
| Buy them from abroad<br>and bring them back | 11.4 (8.9-13.9) | 12.5 (6.3-18.6) | 7.7 (2.6-12.9) | 0.686 |
| Friends/family bring | 0.2 (0.0-0.7) | 0.0 (0.0-0.0) | 1.3 (0.0-3.8) | 0.324 |
| from abroad |  |  |  |  |

<sup>1</sup>All values for Cramer's V were $\leq 0.10$ .

Table 5 shows the sources of purchasing cigarettes among those who smoked menthol cigarettes in England, Scotland, and Wales (unweighted data in supplement, Table S6). All differences between nations were non-significant. The main sources of purchase for all nations were small shops (i.e., newsagent, off license, or corner shop) and supermarkets.

### Sensitivity analyses

Additional sensitivity analyses including all participants who reported that they smoked flavoured cigarettes (instead of only those who reported smoking menthol cigarettes) and the prevalence among all adults (i.e., the proportion of individuals smoking menthol cigarettes among all participants) are presented in the supplement (Tables S7-S13 and Figures S1-S4). The first sensitivity analysis including all people who smoked any flavoured cigarettes did not show meaningful differences compared to the main analysis. In the second sensitivity analysis investigating prevalence of smoking menthol cigarettes as a proportion of all adults (or all 18-to-24-year-olds, respectively), the difference in the prevalence between all adults and 18-to-24-year-olds became more pronounced (see Table S12 and Figure S3). Also, the decrease in prevalence was greater among 18-to-24-year-olds when calculating the prevalence among all participants (PR=0.64, 95% CI: 0.53-0.77, modelled prevalence for Q4 2020: 5.7%, and for Q1 2023: 3.7%) compared to the prevalence among only those who smoked (PR=0.75, 95% CI: 0.63-0.89, modelled prevalence for Q4 2020: 25.7%, and for Q1 2023: 19.4%). For the between-nation comparison, the results for prevalence among all adults and prevalence among those who smoked were comparable.

## DISCUSSION

### Summary of findings

Despite being banned in 2020, one million adults continue to smoke menthol cigarettes in Great Britain. The prevalence of menthol cigarette smoking only decreased slightly and non-significantly among adults who smoke, from 16% at the end of 2020 to 14% at the beginning of 2023. During the same period, the prevalence among 18-to-24-year-olds dropped by a quarter from 26% to 19%. These figures show that despite the ban, menthol cigarette smoking remains common among people who smoke in Great Britain, used by roughly 1 in 7 adults who smoke and 1 in 5 among young adults who smoke. The only nation with a substantial decline in the prevalence was Wales, where it fell by two-thirds from 23% to 8%. Compared with England and Scotland, Wales started off with a higher prevalence, but by the beginning of 2023 it had the lowest prevalence. In contrast, the prevalence in England and Scotland remained relatively stable throughout the period, but in Scotland it was consistently lower than in England.

It is unclear why the trend in Wales differed from the other two nations. Potential contributors could include differences in government approaches and differences in purchasing sources. Since our data on purchasing sources did not differentiate whether people bought menthol cigarettes or accessories to mentholate their cigarettes, we may have missed differences in purchasing sources by nation which are specific to menthol cigarettes. Another explanation could be that the tobacco industry focused its marketing tactics for legal menthol accessories on larger urban areas, mainly in England, rather than less populous localities found in Wales. A further, more unlikely explanation is that people who smoked menthol cigarettes in Wales at the start of the study period were more likely to have quit smoking altogether. When comparing England and Scotland, a potential explanation for the lower overall prevalence in Scotland could be the display ban for tobacco-related accessories in Scotland. Whereas tobacco products (cigarettes, loose tobacco, cigars, etc.) are subject to a display ban in all three nations, only Scotland bans tobacco product accessories from being visible at the point of sale. This difference in legislation could mean that people living in Scotland may be less aware of menthol-flavoured accessories.

There were no noteworthy differences in purchasing sources between menthol and non-flavoured cigarettes; small shops and supermarkets were the most popular places to buy both kinds of cigarettes. There was no evidence that people smoking menthol cigarettes were particularly likely to obtain these from abroad or through illicit sources. Rather, people continued to buy menthol cigarettes in regular shops. This indicates that they either purchased menthol-flavoured accessories or cigarettes they perceived to be mentholated, but which are not labelled as such by the manufacturer.

### Comparison to existing literature

Canada was one of the first countries to introduce a menthol ban and, in contrast to the UK and the European Union, it completely prohibited menthol and its analogues and derivatives in cigarettes nationwide from 2017 [28]. In a pre-post ban comparison, only 20% of people who reported smoking menthol cigarettes before the ban continued doing so afterwards, while 59% switched to non-menthol cigarettes and 22% stopped smoking entirely [29]. In a Dutch longitudinal study, people who smoked menthol cigarettes before the ban there (in March 2020) were asked again about their cigarette smoking behaviour in June-July 2021 [30]. The results showed that 33% of them reported still smoking menthol cigarettes as their usual brand, 40% switching to non-menthol cigarettes, 1% smoking cigarettes with unknown flavour, and 26% having quit smoking. These two studies demonstrate that in other countries people also continued smoking menthol cigarettes despite the ban.

Data from Canada also refute the hypothesis that menthol cigarette bans lead to a surge in illicit purchases [31]. Similarly, Laverty et al. [32] found that the implementation of standardised packaging in the European Union did not lead to an increase in the availability of illicit cigarettes. These findings strengthen the alternative hypothesis that people who continue menthol cigarette smoking after the ban either buy menthol-flavoured accessories or cigarettes that are perceived as being mentholated by consumers but not labelled as such by the manufacturers [13, 29, 30, 33, 34].

### Limitations

This study has several limitations. It is cross-sectional and it is based on self-reported data. No data were available on menthol cigarette smoking prevalence and the purchasing sources for those smoking menthol cigarettes prior to the ban because the question about menthol smoking was only included in the survey after the legislation came into force. The purchasing sources are not specific to where menthol cigarettes were purchased (i.e., they refer to any cigarettes) and it is difficult to clearly classify some of the sources as licit or illicit (e.g., pub behind the bar). One should also note that all the sources of purchase for menthol cigarettes, including supermarkets, could in theory be illicit given the menthol ban in the UK. Further, the question about menthol cigarette smoking in the survey does not provide information about which type of menthol cigarette participants were smoking.

## CONCLUSIONS

Despite the ban, menthol cigarette smoking prevalence remains relatively high among adults in Great Britain as a whole. The cigarette purchasing sources reported by people who smoked menthol cigarettes indicate that mostly people bought these in regular shops and there was no noteworthy difference to those who only smoked non-flavoured cigarettes. Taking these results together with findings from previous studies, it appears that for an effective ban on menthol cigarettes, legislators should close loopholes, such as strictly prohibiting menthol and all its analogues and derivatives in any tobacco-related products. Additionally, better controls might be required to ensure that manufacturers follow these rules.

## ETHICAL APPROVAL

The University College London Ethics Committee granted ethical approval for the Smoking and Alcohol Toolkit Study (ID 0498/001).

## DATA AVAILABILITY

Data are available upon request. The command syntax for the statistical analyses is available on the Open Science Framework (https://osf.io/s8mjr/).

