## Supplementary material for "Smoking prevalence and purchasing of menthol cigarettes since the menthol flavour ban in Great Britain: a population-based survey between 2020 and 2023"

### 1. Missing values

Table S1: Proportion of data missing for each variable (N = 67,746).

| *Variable* | *Missing values, n (%)* |
|---|---:|
| Age | 0 (0) |
| Gender | 110 (0.2) |
| Nation | 0 (0) |
| Social grade | 0 (0) |
| Cigarette smoking | 563 (0.8) |
| Flavoured cigarettes (cigarette smoking n = 9,195) | 208 (2.3) |
| Source of purchase (cigarette smoking n = 6,757) * | 191 (2.8) |

* Question about source of purchase not asked in England in May, June, August, September, November, and December 2022, and in February and March 2023. Therefore, these months were excluded for this question.

### 2. Sensitivity analysis: unweighted

Table S2: Characteristics of participants, unweighted vs. weighted (N = 66,868).

| *Characteristic* | *Unweighted* | *Weighted* |
|---|---|---|
| Age, median (IQR) | 52 (35-66) | 49 (33-63) |
| Gender, n (%) | | |
| Female | 34108 (51.0) | 33918 (50.8) |
| Male | 32360 (48.4) | 32427 (48.6) |
| Non-binary | 400 (0.6) | 400 (0.6) |
| Social grade, n (%) | | |
| ABC1 | 43808 (65.5) | 37341 (55.9) |
| C2DE | 23060 (34.5) | 29404 (44.1) |
| Nation, n (%) | | |
| England | 47641 (71.2) | 57628 (86.3) |
| Scotland | 12442 (18.6) | 5842 (8.8) |
| Wales | 6785 (10.1) | 3275 (4.9) |
| Smoking cigarettes, n (%) | 8965 (13.4) | 9773 (14.6) |

Table S3: Prevalence of menthol cigarette smoking in different population groups who smoke cigarettes (unmodelled, weighted).

| *Quarter (Q)* | *Great Britain, all adults, % (95% CI)* | *Great Britain, 18-24-year-olds, % (95% CI)* | *England, all adults, % (95% CI)* | *Scotland, all adults, % (95% CI)* | *Wales, all adults, % (95% CI)* |
|---|---|---|---|---|---|
| Q4 2020 | 17.5 (14.8-20.3) | 24.5 (16.7-32.3) | 17.7 (14.6-20.8) | 11.0 (5.0-17.0) | 23.5 (13.9-33.2) |
| Q1 2021 | 16.7 (14.0-19.4) | 30.4 (21.4-39.3) | 17.0 (14.0-20.0) | 11.8 (7.1-16.5) | 19.6 (10.8-28.4) |
| Q2 2021 | 12.7 (10.2-15.2) | 15.9 (9.6-22.3) | 12.0 (9.3-14.8) | 15.3 (9.5-21.0) | 22.0 (11.0-32.9) |
| Q3 2021 | 14.9 (12.2-17.6) | 14.4 (8.1-20.7) | 15.7 (12.6-18.8) | 9.4 (4.9-13.9) | 10.8 (3.0-18.7) |
| Q4 2021 | 16.2 (13.5-19.0) | 18.9 (11.0-26.9) | 16.6 (13.4-19.7) | 9.8 (5.4-14.2) | 24.4 (13.5-35.3) |
| Q1 2022 | 18.7 (15.7-21.7) | 26.7 (17.9-35.5) | 20.0 (16.6-23.4) | 10.2 (5.2-15.1) | 11.6 (5.1-18.0) |
| Q2 2022 | 13.9 (11.2-16.6) | 15.8 (9.4-22.2) | 14.4 (11.3-17.4) | 10.9 (5.2-16.7) | 10.9 (4.1-17.7) |
| Q3 2022 | 14.0 (11.5-16.6) | 15.3 (8.8-21.8) | 13.8 (11.0-16.6) | 15.9 (9.7-22.0) | 15.5 (7.9-23.1) |
| Q4 2022 | 15.0 (12.2-17.7) | 15.9 (9.2-22.7) | 15.6 (12.5-18.7) | 11.6 (6.4-16.9) | 7.5 (2.2-12.8) |
| Q1 2023 | 13.0 (10.4-15.6) | 22.4 (13.6-31.1) | 13.6 (10.7-16.6) | 8.8 (3.9-13.8) | 7.7 (1.1-14.4) |

Table S4: Prevalence of menthol cigarette smoking among people who smoke cigarettes (unmodelled, unweighted).

| *Quarter (Q)* | *Great Britain, all adults, % (95% CI)* | *Great Britain, 18-24-year-olds, % (95% CI)* | *England, all adults, % (95% CI)* | *Scotland, all adults, % (95% CI)* | *Wales, all adults, % (95% CI)* |
|---|---|---|---|---|---|
| Q4 2020 | 17.6 (15.1-20.1) | 25.4 (18.1-32.6) | 18.3 (15.4-21.3) | 10.3 (5.1-15.5) | 23.3 (14.4-32.2) |
| Q1 2021 | 16.7 (14.3-19.0) | 29.0 (21.1-36.9) | 17.5 (14.6-20.4) | 12.6 (7.9-17.4) | 18.9 (10.9-27) |
| Q2 2021 | 13.0 (10.8-15.2) | 20.0 (13.3-26.7) | 11.6 (9.2-14.1) | 16.0 (10.3-21.8) | 18.4 (9.5-27.3) |
| Q3 2021 | 13.7 (11.4-15.9) | 16.9 (10.4-23.5) | 15.1 (12.4-17.9) | 9.9 (5.4-14.4) | 10.0 (3.3-16.7) |
| Q4 2021 | 15.6 (13.2-18.0) | 18.9 (11.5-26.3) | 15.8 (13.0-18.6) | 11.4 (6.6-16.3) | 22.2 (13.0-31.5) |
| Q1 2022 | 16.6 (14.2-19.0) | 24.6 (17.1-32.1) | 19.2 (16.1-22.2) | 9.9 (5.5-14.3) | 12.5 (6.0-19.0) |
| Q2 2022 | 12.9 (10.7-15.2) | 17.8 (11.2-24.3) | 13.7 (11.0-16.4) | 10.8 (5.6-16) | 11.0 (4.4-17.5) |
| Q3 2022 | 14.9 (12.6-17.2) | 16.1 (10.0-22.2) | 14.5 (11.8-17.2) | 15 (9.7-20.4) | 17.4 (9.5-25.3) |
| Q4 2022 | 14.6 (12.2-16.9) | 18.1 (11.3-24.9) | 15.9 (13.0-18.8) | 12.4 (7.3-17.6) | 9.2 (3.0-15.4) |
| Q1 2023 | 12.3 (10.1-14.6) | 20.5 (12.9-28.1) | 13.6 (10.9-16.3) | 9.6 (4.6-14.6) | 7.2 (1.5-12.9) |

Table S5: Sources of cigarette purchases (not mutually exclusive) among those who smoke cigarettes (unweighted)

| ***Source of purchase*** | ***Among people smoking menthol cigarettes, n (%)*** | ***Among people smoking non-flavoured cigarettes, n (%)*** | ***P-value[1]*** |
|---|---|---|---|
| Newsagent\Off licence\Corner shop | 716 (71.3) | 3755 (68.6) | 0.096 |
| Petrol garage shop | 417 (41.5) | 2140 (39.1) | 0.157 |
| Supermarket | 725 (72.2) | 4007 (73.2) | 0.535 |
| Cash and Carry | 57 (5.7) | 256 (4.7) | 0.201 |
| Internet | 25 (2.5) | 145 (2.6) | 0.855 |
| Bar in pub | 19 (1.9) | 69 (1.3) | 0.150 |
| Other sources | 6 (0.6) | 57 (1.0) | 0.325 |
| Illicit | | | |
| Newsagent\Off license\Corner shop – "under the counter" | 75 (7.5) | 294 (5.4) | 0.01 |
| Friends | 54 (5.4) | 272 (5.0) | 0.641 |
| Trusted local | 34 (3.4) | 176 (3.2) | 0.854 |
| Person in pub | 24 (2.4) | 86 (1.6) | 0.087 |
| Person on the street | 21 (2.1) | 113 (2.1) | 1.000 |
| Vending machine pub | 9 (0.9) | 41 (0.7) | 0.769 |
| Cross-border | | | |
| Buy them from abroad and bring them back | 108 (10.8) | 539 (9.8) | 0.409 |
| Friends/family bring from abroad | 2 (0.2) | 7 (0.1) | 0.923 |

[1]All values for Cramer's V were ≤0.10.

Table S6: Sources of cigarette purchases (not mutually exclusive) by nation (unweighted).

| ***Source of purchase*** | ***Among people smoking menthol cigarettes in England, n (%)*** | ***Among people smoking menthol cigarettes in Scotland, n (%)*** | ***Among people smoking menthol cigarettes in Wales, n (%)*** | ***P-value[1]*** |
|---|---|---|---|---|
| Newsagent/Off licence/Corner shop | 546 (71.9) | 98 (69.0) | 72 (69.9) | 0.737 |
| Petrol garage shop | 315 (41.5) | 56 (39.4) | 46 (44.7) | 0.715 |
| Supermarket | 545 (71.8) | 102 (71.8) | 78 (75.7) | 0.702 |
| Cash and Carry | 44 (5.8) | 9 (6.3) | 4 (3.9) | 0.686 |
| Internet | 17 (2.2) | 6 (4.2) | 2 (1.9) | 0.353 |
| Bar in pub | 16 (2.1) | 2 (1.4) | 1 (1.0) | 0.657 |
| Other sources | 5 (0.7) | 1 (0.7) | 0 (0.0) | 0.722 |
| Illicit | | | | |
| Newsagent/Off license/ Corner shop – “under the counter” | 60 (7.9) | 5 (3.5) | 10 (9.7) | 0.125 |
| Friends | 44 (5.8) | 3 (2.1) | 7 (6.8) | 0.162 |
| Trusted local | 29 (3.8) | 0 (0.0) | 5 (4.9) | 0.048 |
| Person in pub | 20 (2.6) | 3 (2.1) | 1 (1.0) | 0.568 |
| Person on the street | 16 (2.1) | 3 (2.1) | 2 (1.9) | 0.994 |
| Vending machine pub | 8 (1.1) | 1 (0.7) | 0 (0.0) | 0.548 |
| Cross-border | | | | |
| Buy them from abroad and bring them back | 83 (10.9) | 16 (11.3) | 9 (8.7) | 0.778 |
| Friends/family bring from abroad | 1 (0.1) | 0 (0.0) | 1 (1.0) | 0.170 |

[1]All values for Cramer’s V were ≤0.10.

#### 3. Sensitivity analysis: using all flavoured cigarettes instead of only menthol cigarettes

Table S7: Prevalence of flavoured cigarette smoking in different population groups who smoke cigarettes (unmodelled, weighted).

| *Quarter (Q)* | *Great Britain, all adults, % (95% CI)* | *Great Britain, 18-24-year-olds, % (95% CI)* | *England, all adults, % (95% CI)* | *Scotland, all adults, % (95% CI)* | *Wales, all adults, % (95% CI)* |
|---|---|---|---|---|---|
| Q4 2020 | 20.1 (17.1-23.1) | 25.7 (17.8-33.6) | 20.5 (17.2-23.8) | 11.6 (5.5-17.7) | 25.0 (15.3-34.8) |
| Q1 2021 | 18.8 (16.0-21.6) | 34.8 (25.6-43.9) | 18.9 (15.7-22.0) | 16.7 (11.0-22.4) | 21.9 (12.9-30.9) |
| Q2 2021 | 15.7 (13.0-18.4) | 19.7 (12.6-26.8) | 15.1 (12.1-18.1) | 18.2 (11.5-24.9) | 24.4 (13.2-35.5) |
| Q3 2021 | 17.0 (14.2-19.8) | 17.1 (10.3-23.9) | 17.8 (14.6-21.0) | 11.2 (6.3-16.2) | 14.0 (5.1-22.8) |
| Q4 2021 | 18.2 (15.3-21.1) | 24.1 (15.2-33.1) | 18.5 (15.2-21.8) | 11.6 (6.7-16.5) | 25.5 (14.5-36.5) |
| Q1 2022 | 21.4 (18.2-24.6) | 30.8 (21.6-40.0) | 23.0 (19.3-26.6) | 12.0 (6.8-17.2) | 12.1 (5.6-18.6) |
| Q2 2022 | 15.6 (12.8-18.4) | 17.1 (10.5-23.7) | 15.9 (12.7-19.0) | 13.5 (7.3-19.8) | 14.1 (6.6-21.7) |
| Q3 2022 | 16.9 (14.2-19.7) | 18.8 (11.6-26.0) | 16.8 (13.7-19.9) | 17.6 (11.3-23.8) | 18.2 (10.1-26.3) |
| Q4 2022 | 17.6 (14.7-20.6) | 16.8 (10.0-23.7) | 18.2 (14.9-21.5) | 13.5 (7.7-19.3) | 12.8 (5.7-20.0) |
| Q1 2023 | 14.3 (11.6-17.0) | 26.4 (17.1-35.7) | 15.0 (12.0-18.1) | 8.8 (3.9-13.8) | 10.1 (2.3-17.9) |

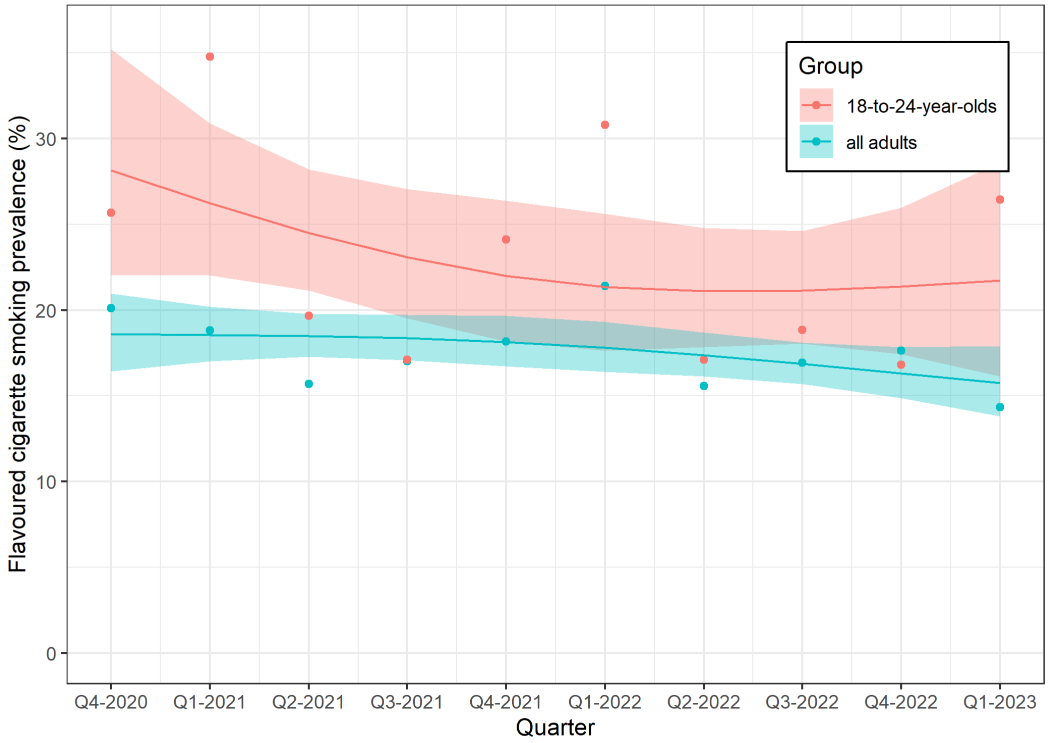


Figure S1: Weighted prevalence of smoking flavoured cigarettes among all adults who smoke cigarettes and those aged 18-24 years in Great Britain over time. Lines and shaded bands represent point estimates and 95% compatibility intervals, respectively, from logistic regression with time modelled with restricted cubic splines (3 knots). The points represent unmodelled data.

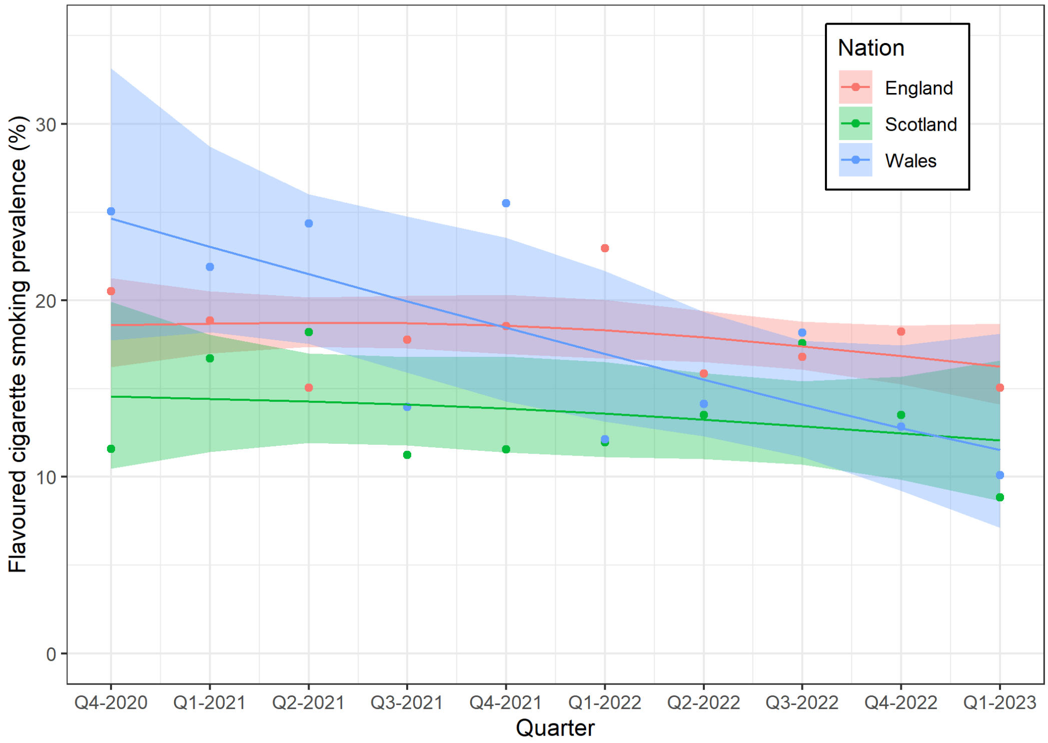


Figure S2: Weighted prevalence of smoking flavoured cigarettes among adults smoking cigarettes in England, Scotland, and Wales over time. Lines and shaded bands represent point estimates and 95% compatibility intervals, respectively, from logistic regression with time modelled with restricted cubic splines (3 knots). The points represent unmodelled data.

Table S8: Modelled prevalence and prevalence ratios comparing first (Q4 2020) to last quarter (Q1 2023) (weighted).

| ***Sample*** | ***Menthol flavour*** | | | ***Any flavour*** | | |
|---|---|---|---|---|---|---|
| | Q4 2020, % (95% CI) | Q1 2023, % (95% CI) | PR (95% CI) | Q4 2020, % (95% CI) | Q1 2023, % (95% CI) | PR (95% CI) |
| All adults | 16.2 (14.1-18.4) | 13.7 (11.9-15.8) | 0.85 (0.71-1.01) | 18.6 (16.4-21.0) | 15.7 (13.8-17.9) | 0.85 (0.72-0.99) |
| 18-to-24-year-olds | 25.7 (19.7-32.8) | 19.4 (11.9-26.0) | 0.75 (0.63-0.89) | 28.1 (22.0-35.2) | 21.7 (13.8-28.6) | 0.77 (0.61-0.98) |
| Adults in England | 16.2 (13.9-18.7) | 14.2 (12.2-16.5) | 0.88 (0.72-1.06) | 18.6 (16.2-21.3) | 16.3 (14.1-18.7) | 0.87 (0.73-1.05) |
| Adults in Scotland | 12.0 (8.3-17.0) | 11.3 (8.0-15.8) | 0.94 (0.59-1.53) | 14.6 (10.5-19.9) | 12.1 (8.6-16.6) | 0.83 (0.53-1.31) |
| Adults in Wales | 22.5 (15.8-31.0) | 8.1 (4.7-13.7) | 0.36 (0.19-0.62) | 24.6 (17.8-33.1) | 11.5 (7.1-18.1) | 0.47 (0.27-0.79) |
| Adults in Scotland vs. England | NA | NA | 1.07 (0.64-1.80) | NA | NA | 0.95 (0.58-1.55) |
| Adults in Wales vs. England | NA | NA | 0.41 (0.21-0.75) | NA | NA | 0.54 (0.30-0.93) |

Table S9: Purchase sources (not mutually exclusive) among those smoking flavoured vs non-flavoured cigarettes (weighted).

| *Source of purchase* | *Among people smoking flavoured cigarettes, % (95% CI)* | *Among people smoking non-flavoured cigarettes, % (95% CI)* | *P-value[1]* |
|---|---|---|---|
| Newsagent\Off licence\Corner shop | 72.4 (69.4-75.3) | 69.8 (68.4-71.1) | 0.078 |
| Petrol garage shop | 42.7 (39.4-46) | 40.9 (39.3-42.4) | 0.251 |
| Supermarket | 72.3 (69.3-75.2) | 72.2 (70.8-73.5) | 0.935 |
| Cash and Carry | 6.5 (4.7-8.2) | 5.3 (4.5-6.0) | 0.104 |
| Internet | 2.6 (1.5-3.7) | 2.7 (2.2-3.2) | 0.857 |
| Bar in pub | 1.8 (0.9-2.8) | 1.3 (0.9-1.6) | 0.157 |
| Other sources | 0 (0.0-0.0) | 0.9 (0.7-1.2) | 0.127 |
| Illicit | | | |
| Newsagent\Off license\Corner shop – "under the counter" | 8.1 (6.2-10.0) | 6.0 (5.3-6.8) | 0.009 |
| Friends | 5.8 (4.2-7.3) | 5.5 (4.8-6.3) | 0.764 |
| Trusted local | 3.6 (2.3-4.9) | 3.5 (2.9-4.1) | 0.882 |
| Person in pub | 2.8 (1.6-3.9) | 1.8 (1.4-2.2) | 0.027 |
| Person on the street | 2.4 (1.4-3.4) | 2.3 (1.8-2.8) | 0.791 |
| Vending machine pub | 1.1 (0.3-1.9) | 0.9 (0.6-1.2) | 0.452 |
| Cross-border | | | |
| Buy them from abroad and bring them back | 11.3 (9.1-13.5) | 9.8 (8.9-10.7) | 0.114 |
| Friends/family bring from abroad | 0.2 (0.0-0.6) | 0.1 (0.0-0.2) | 0.327 |

[1]All values for Cramer's V were ≤0.10.

Table S10: Type of purchase (not mutually exclusive) among those smoking flavoured cigarettes by nation (weighted).

| *Type of purchase* | *Among people smoking flavoured cigarettes, % (95% CI)* | *Among people smoking non-flavoured cigarettes, % (95% CI)* |
|---|---|---|
| Licit | 93.6 (92.0-95.2) | 93.5 (92.7-94.2) |
| Illicit | 14.7 (12.3-17.1) | 12.5 (11.4-13.5) |
| Cross-border | 11.5 (9.4-13.7) | 9.9 (9.0-10.8) |

Table S11: Purchase sources (not mutually exclusive) among those smoking flavoured cigarettes by nation (weighted).

| ***Source of purchase*** | ***Among people smoking flavoured cigarettes in England, % (95% CI)*** | ***Among people smoking menthol cigarettes in Scotland, % (95% CI)*** | ***Among people smoking menthol cigarettes in Wales, % (95% CI)*** | ***P-value[1]*** |
|---|---|---|---|---|
| Newsagent/Off licence/Corner shop | 72.9 (69.7-76.0) | 67.3 (59.4-75.3) | 70.4 (61.3-79.5) | 0.561 |
| Petrol garage shop | 42.2 (38.6-45.8) | 42.4 (34.1-50.7) | 47.4 (37.4-57.4) | 0.733 |
| Supermarket | 72.0 (68.7-75.2) | 72.1 (64.6-79.7) | 76.6 (68.2-84.9) | 0.741 |
| Cash and Carry | 6.6 (4.8-8.5) | 7.6 (2.9-12.2) | 4.2 (0.1-8.2) | 0.709 |
| Internet | 2.7 (1.6-3.9) | 3.3 (0.8-5.7) | 3.1 (0.0-6.8) | 0.956 |
| Bar in pub | 2.1 (1.1-3.2) | 1.3 (0.0-3.4) | 0.6 (0.0-1.8) | 0.638 |
| Other sources | 0.5 (0.0-1.0) | 0.4 (0.0-1.1) | 0.0 (0.0-0.0) | 0.844 |
| Illicit | | | | |
| Newsagent/Off license/Corner shop – “under the counter” | 8.5 (6.4-10.6) | 5.3 (1.5-9.2) | 9.5 (3.4-15.6) | 0.607 |
| Friends | 6.2 (4.4-7.9) | 6.1 (1.3-10.9) | 4.6 (1.1-8.2) | 0.888 |
| Trusted local | 3.9 (2.5-5.3) | 2.8 (0.0-6.3) | 5.1 (0.7-9.5) | 0.801 |
| Person in pub | 2.9 (1.7-4.2) | 4.9 (0.5-9.4) | 1.7 (0.0-4.9) | 0.524 |
| Person on the street | 2.4 (1.4-3.5) | 4.9 (0.6-9.1) | 3.1 (0.0-6.9) | 0.444 |
| Vending machine pub | 1.1 (0.3-1.9) | 1.1 (0.0-3.2) | 1.7 (0.0-4.9) | 0.925 |
| Cross-border | | | | |
| Buy them from abroad and bring them back | 11.5 (9.2-13.9) | 14.1 (8.0-20.2) | 8.9 (3.4-14.4) | 0.646 |
| Friends/family bring from abroad | 0.2 (0.0-0.6) | 0.0 (0.0-0.0) | 1.1 (0.0-3.3) | 0.323 |

[1]All values for Cramer’s V were ≤0.10.

**4. Sensitivity analysis: prevalence of smoking menthol cigarettes as a proportion of all adults (or all 18-to-24-year-olds)**

Table S12: Prevalence of cigarette smoking among all participants in different population groups (unmodelled, weighted).

| ***Quarter (Q)*** | ***Great Britain, all adults, % (95% CI)*** | ***Great Britain, 18-24-year-olds, % (95% CI)*** | ***England, all adults, % (95% CI)*** | ***Scotland, all adults, % (95% CI)*** | ***Wales, all adults, % (95% CI)*** |
|---|---|---|---|---|---|
| Q4 2020 | 2.6 (2.1-3.0) | 5.5 (3.5-7.4) | 2.6 (2.1-3.1) | 1.3 (0.6-2.1) | 3.7 (2.0-5.4) |
| Q1 2021 | 2.5 (2.1-2.9) | 6.0 (3.9-8.1) | 2.6 (2.1-3.1) | 1.8 (1.0-2.5) | 2.8 (1.4-4.1) |
| Q2 2021 | 1.9 (1.5-2.3) | 4.1 (2.4-5.8) | 1.9 (1.4-2.3) | 2.2 (1.3-3.1) | 2.7 (1.2-4.2) |
| Q3 2021 | 2.2 (1.7-2.6) | 2.8 (1.5-4.1) | 2.3 (1.8-2.7) | 1.4 (0.7-2.1) | 1.6 (0.4-2.8) |
| Q4 2021 | 2.4 (1.9-2.8) | 3.5 (1.9-5.1) | 2.4 (1.9-2.8) | 1.7 (0.9-2.5) | 3.5 (1.7-5.3) |
| Q1 2022 | 2.7 (2.3-3.2) | 5.1 (3.2-7.1) | 2.9 (2.4-3.5) | 1.5 (0.7-2.2) | 2.0 (0.8-3.1) |
| Q2 2022 | 2.0 (1.6-2.4) | 2.9 (1.7-4.2) | 2.1 (1.6-2.5) | 1.5 (0.6-2.3) | 1.7 (0.6-2.9) |
| Q3 2022 | 2.1 (1.7-2.5) | 3.2 (1.7-4.6) | 2.1 (1.7-2.6) | 2.1 (1.2-3.0) | 2.2 (1.1-3.4) |
| Q4 2022 | 2.1 (1.7-2.6) | 2.8 (1.6-4.1) | 2.3 (1.8-2.8) | 1.5 (0.8-2.2) | 1.0 (0.3-1.6) |
| Q1 2023 | 1.8 (1.4-2.2) | 4.2 (2.4-6.0) | 1.9 (1.5-2.4) | 1.1 (0.5-1.7) | 1.2 (0.1-2.2) |

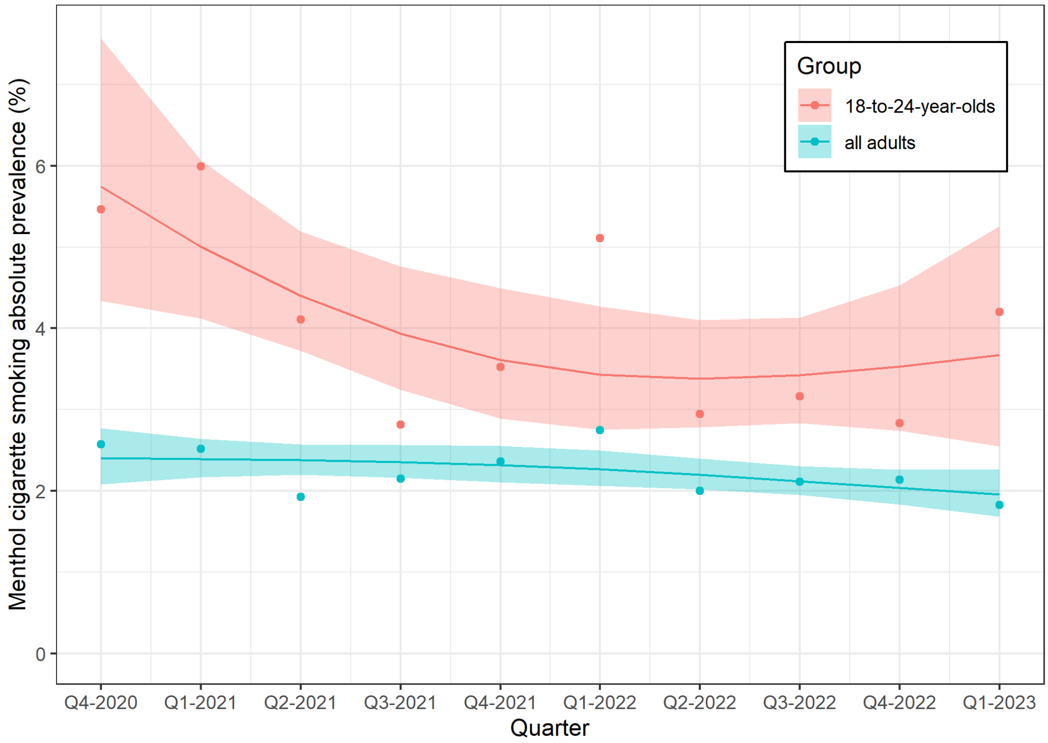


Figure S3: Weighted prevalence of smoking menthol cigarettes among all adults and those aged 18-24 years in Great Britain over time. Lines and shaded bands represent point estimates and 95% compatibility intervals, respectively, from logistic regression with time modelled with restricted cubic splines (3 knots). The points represent unmodelled data.

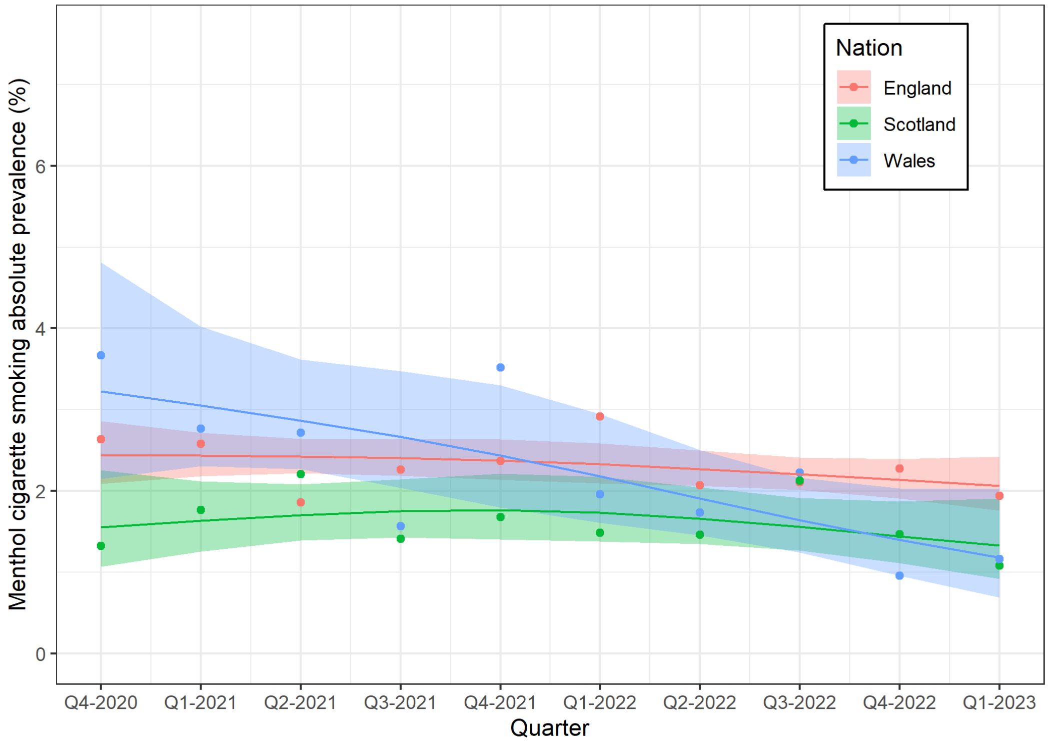


Figure S4: Weighted prevalence of smoking flavoured cigarettes among adults in England, Scotland, and Wales over time. Lines and shaded bands represent point estimates and 95% compatibility intervals, respectively, from logistic regression with time modelled with restricted cubic splines (3 knots). The points represent unmodelled data.

Table S13: Modelled prevalence and prevalence ratios comparing first (Q4 2020) to last quarter (Q1 2023) (weighted), comparing prevalence (i.e., among all who smoke) to prevalence among all participants.

| ***Sample*** | ***Prevalence among those who smoke*** | | | ***Prevalence among all participants*** | | |
|---|---|---|---|---|---|---|
| | Q4 2020, % (95% CI) | Q1 2023, % (95% CI) | PR (95% CI) | Q4 2020, % | Q1 2023, % (95% CI) | PR (95% CI) |
| All adults | 16.2 (14.1-18.4) | 13.7 (11.9-15.7) | 0.85 (0.71-1.01) | 2.4 (2.1-2.8) | 2.0 (1.7-2.3) | 0.81 (0.68-0.98) |
| 18-to-24-year-olds | 25.7 (19.7-32.8) | 19.4 (14.1-26.0) | 0.75 (0.63-0.90) | 5.7 (4.3-7.6) | 3.7 (1.7-5.3) | 0.64 (0.53-0.77) |
| Adults in England | 6.2 (13.9-18.7) | 14.2 (12.2-16.5) | 0.88 (0.73-1.06) | 2.4 (2.1-2.9) | 2.1 (1.8-2.4) | 0.84 (0.69-1.04) |
| Adults in Scotland | 12.0 (8.3-17.0) | 11.3 (7.9-15.8) | 0.94 (0.58-1.50) | 1.6 (1.1-2.3) | 1.3 (0.9-1.9) | 0.85 (0.50-1.38) |
| Adults in Wales | 22.5 (15.8-31.0) | 8.1 (4.6-13.7) | 0.36 (0.19-0.62) | 3.2 (2.2-4.8) | 1.2 (0.7-2.0) | 0.37 (0.19-0.68) |
| Adults in Scotland vs. England | NA | NA | 1.07 (0.63-1.79) | NA | NA | 1.00 (0.57-1.66) |
| Adults in Wales vs. England | NA | NA | 0.41 (0.20-0.71) | NA | NA | 0.43 (0.22-0.82) |
